# Current landscape of disparity-focused research: a bibliometric analysis of 260 research articles

**DOI:** 10.1101/2023.03.05.23286652

**Authors:** Anushka Walia, Alyson Haslam, Jordan Tuia, Milos Milijković, Vinay Prasad

**Author notes:** **Corresponding author:** Alyson Haslam, PhD, Department of Epidemiology and Biostatistics, UCSF Mission Bay Campus | Mission Hall: Global Health & Clinical Sciences Building | 550 16th St, 2nd Fl, San Francisco, CA 94158.

## Abstract

The health disparities research field is rapidly evolving, but not all populations may be adequately represented in research on disparities. This study characterizes the current state of disparities literature and identifies trends, gaps, and opportunities in the field through bibliometric analysis. 260 articles published between January 2020 and August 2022, of which 95% focused on health disparities, were included in the analysis. 67% of studies investigated disparities across racial or ethnic groups, and all 9 articles on physician employment investigated disparities in gender. The majority of studies focused on adults, with 71% of studies having mean or median ages of subjects between 40 and 80 years. Articles on cancer and cardiovascular conditions comprised 34% of health-related disparity studies. Our results highlight a need to study disparities in pediatric and young adult populations. Such research can identify targets of early intervention that may improve long-term health and reduce disparities presenting in late adulthood.

## Introduction

Disparities exist across all domains of the healthcare system, from health outcomes to access to care to career opportunities. Health disparities research output has grown since 1990, with approximately 50,000 papers published in the field by 2016.[1] These studies have revealed inequities across a broad range of socio-demographic variables including race, ethnicity, sex, socioeconomic status, sexual orientation, and geography. The COVID-19 crisis has disproportionally burdened racial and ethnic minority populations, highlighting deep structural inequalities that have existed for decades.[2] Research characterizing disparities and implementing interventions to address them are more vital than ever. Publications have recently acknowledged the critical responsibility of scientific journals in dismantling health disparities and the body of disparities literature is expected to grow.[3]

As disparities may manifest in all aspects of medicine, it is important to study a variety of health conditions, sources of disparity, and populations. However, not all groups may be equally represented in disparities research. For instance, while disparities in both pediatric and adult health have been well-documented, pediatric health disparity research has traditionally received less funding than research on adults and remains more limited in scope.[4] Yet, studies on pediatric disparities are essential, particularly because childhood disparities can be determinants of health outcomes in adulthood. Such research may identify targets of early intervention to bridge disparities that occur later in life, and interventional approaches in children may differ from those applied to adults.[5]

It is necessary to understand the publication patterns of disparities research to identify potential gaps in the field. To our knowledge, there has been no attempt to systematically survey the landscape of disparities research in recent years. Prior bibliometric analyses of disparities research were conducted prior to 2016, included only top-cited papers, or focused on specific health disparities, countries, or areas of medicine. [1] [6] [7] In addition, no prior study has examined the ages of subjects included in disparities research to identify underrepresented age groups. In this study, we perform a systematic analysis of research articles on disparities to identify current subject areas, trends, and imbalances in the disparities field.

## Methods

### Search strategy

We searched PubMed for articles containing the terms “disparity[Title] OR disparities[Title]” from January 2020 to August 2022 (Fig 1). Articles that fell under the categories classical article, clinical study, clinical trial, comparative study, controlled clinical trial, multicenter study, observational study, pragmatic clinical trial, randomized controlled trial, and validation study were considered for inclusion. We excluded commentaries, reviews, study protocols, and animal studies. All studies from 2022 and the first 100 articles (sorted by best match) from each of the years 2021 and 2020 were included, resulting in 260 total articles that were further analyzed.

**Fig 1.**
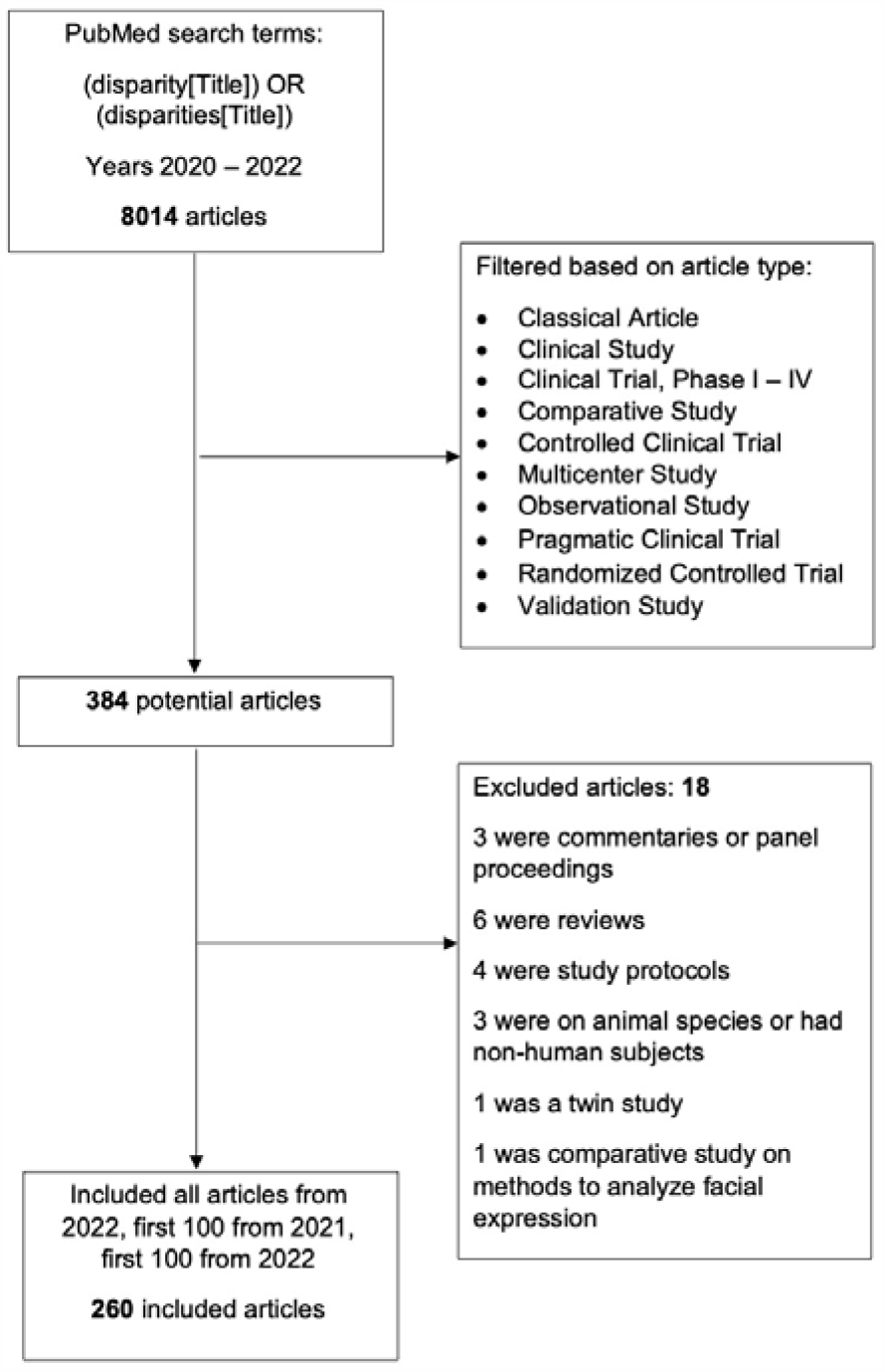
Flowchart describing literature search strategy

### Data extraction

Studies were broadly coded by disparity category: health-related, employment-related, or education-related. For each health-related study, we extracted the types of disparities examined: “clinical outcomes” (e.g., mortality, survival, surgical complications), “treatment or care access” (e.g., treatment modality, linkage to care, time to surgery, vaccine access), “disease/condition risk or rate,” “disease/condition characteristics” (e.g., presence of symptoms, cancer stage at diagnosis), etc. (detailed in Table 1). Studies that examined multiple types of disparities or disparities other than those listed were classified as “multiple/other.” Health-related studies were further coded based on the disease or condition studied. Articles on both COVID-19 and another condition (e.g., COVID-19 and cancer) were coded as “COVID-19 + 1 other condition.” Studies pertaining to cancer were divided based on type of cancer studied.

**Table 1:** Characteristics of published studies examining disparities.

| **Variable** | **Frequency** |
| --- | --- |
| **Total, N** | **260** |
| **Year of publication** |  |
| 2022 (January 1 – August 1) | 60 |
| 2021 | 100 |
| 2020 | 100 |
| **Type of disparity** |  |
| **Health-related** | **248** |
| Clinical outcomes | 66 |
| Treatment or care access | 48 |
| Disease/condition risk or rate | 16 |
| Disease/condition characteristics | 14 |
| Visit type | 8 |
| Hospital management patterns | 10 |
| Screening or testing access | 5 |
| Substance use patterns | 5 |
| Attitudes | 4 |
| Clinical trial access or inclusion | 3 |
| Health status | 4 |
| Policy | 3 |
| Referral patterns | 3 |
| Multiple disparities or other | 59 |
| **Employment** | **9** |
| **School disciplinary action** | **2** |
| **Child welfare investigation** | **1** |
| **Disease or condition of study participants in health-related studies** |  |
| Cancer | 47 |
| Gynecologic | 5 |
| Breast | 5 |
| Lung | 4 |
| Prostate | 4 |
| Hematologic | 3 |
| Head and Neck | 3 |
| Colorectal or rectal | 3 |
| Melanoma | 2 |
| Gastric | 2 |
| Bladder | 2 |
| Glioblastoma | 1 |
| Multiple | 13 |
| Cardiovascular | 38 |
| Obstetrics/Gynecologic | 12 |
| Pulmonary | 11 |
| Diabetes | 8 |
| Orthopedic | 8 |
| Kidney-related or urologic | 8 |
| Psychiatric or substance use | 6 |
| Ophthalmologic | 6 |
| COVID-19 | 6 |
| Gastroenterological | 5 |
| HIV | 4 |
| Autism of intellectual disability | 4 |
| Neurologic | 4 |
| COVID + 1 other condition | 3 |
| Thyroid or parathyroid | 3 |
| Liver | 3 |
| General ED or hospital patients | 8 |
| General primary care or surgical patients | 8 |
| Other | 19 |
| No specific condition or not applicable | 37 |
| **COVID-related (risk, testing, outcomes, policy, etc.)** | **33** |
| **Journal** |  |
| JAMA Network Open | 10 |
| The American Journal of Surgery | 5 |
| Journal of Racial and Ethnic Health Disparities | 4 |
| Laryngoscope | 4 |
| Obstetrics & Gynecology | 4 |
| Surgery | 4 |
| Academic Emergency Medicine | 3 |
| American Journal of Cardiology | 3 |
| Autism Research | 3 |
| Journal of Clinical Oncology | 3 |
| Journal of General Internal Medicine | 3 |
| Journal of Surgical Research | 3 |
| Journal of the American Heart Association | 3 |
| Journal of Vascular Surgery | 3 |
| Pediatrics | 3 |
| Resuscitation | 3 |
| The Annals of Thoracic Surgery | 3 |
| Urology | 3 |
| Other (2 or fewer articles) | 162 |
| **Journal impact factors** |  |
| Low-ranked (<5) | 122 |
| Mid-ranked (5 – 10) | 80 |
| Top-ranked (≥10) | 48 |
| Not available | 10 |

From each article, we extracted the sociodemographic or clinical variables across which disparities were studied and classified them as: “race/ethnicity,” “sex/gender,” “geography,” “rurality,” “socioeconomic status,” “disability,” “insurance,” or “English proficiency.” Studies examining disparities across one or more variables in addition to race or ethnicity (e.g., both race/ethnicity and sex/gender) were given the classification “race/ethnicity + 1 other” or “race/ethnicity + 2 or more.” All other categories of disparities (occupation, neighborhood vulnerability, educational attainment, etc.) were coded as “other.” Studies on two or more non-racial or ethnic disparities (e.g., sex/gender and socioeconomic status) also fell into the “other” category. Classification of disparity type was based on the primary independent variable(s) used for quantitative analysis and not variables that were simply adjusted for in models. For instance, a study that found that socioeconomic status mediated a racial disparity (i.e., adjusting for socioeconomic status eliminated the disparity) was given the classification racial/ethnic.

The median age of subjects in each study was recorded. For studies that did not report median ages, mean ages were used if provided. Many studies provided age brackets and the number of subjects in each bracket rather than a single mean or median value. In these cases, we computed a weighted mean by summing products of the center of each bracket and the fraction of participants in each bracket. Some studies did not provide the lower or upper bounds of first or last age brackets. In these cases, upper or lower bounds were estimated using the following criteria: if the upper bound for the highest age bracket was not provided and the study was conducted in the United States, the average lifespan in the United States (78.8 yrs.) was assumed to be the upper bound. For studies conducted in Australia or China, life expectancies of 82.9 and 76.9 yrs. respectively were used as upper bounds if required. If the lower bound of an age bracket was greater than the average lifespan of the population (e.g., an age bracket of 80+), the upper bound was estimated to be 90, assuming that very few, if any, participants would exceed this age. If a study did not include the lower bound of the first age bracket (e.g., an age bracket of <30) but investigated a condition that predominantly affects adults (e.g., endometrial cancer), the lowest age of participants was assumed to be 18. For studies on pregnancy and maternal health that did not provide youngest or oldest ages of participants, the average age of menarche (12.4 yrs.) and menopause (51 yrs.) were used. For an article on emergency department restraint use, 13 was used as the lowest age of study subjects, assuming that most restraint use occurs in teenage children and older. For a study describing gender disparities in gastroenterology, the youngest age was assumed to be 30 and the oldest 65 (retirement age).

For each study we also recorded the journal title, journal impact factor, and whether or not the study pertained to COVID-19. Journals were divided into three categories based on impact factors: low-ranked (<5), mid-ranked (5-10), and top-ranked (≥10).

## Results

Our search yielded 8014 articles, of which 304 were considered (Fig 1). Our inclusion criteria identified 260 PubMed (2020-2022) articles for the analysis. Characteristics of these articles are described in Table 1. 95% (n=248) of studies described health-related disparities, and the remainder described disparities in employment (3.5%, n=9), school disciplinary action (0.8%, n=2), or child welfare investigation (0.4%, n=1). Among health disparity-related studies, almost half investigated disparities in either clinical outcomes (27%, n=66) or treatment or care access (19%, n=48). The most commonly studied condition was cancer (19%, n = 47), followed by cardiovascular (15%, n=38) and obstetrics/gynecologic (5%, n=12). 33 (13%) of 260 articles related to COVID-19, covering clinical outcomes, screening rates, telehealth use, and other pandemic-related behaviors.

The 260 studies were published in 180 different journals. The journal that contributed the highest number of articles was *JAMA Network Open* (4%, n=10), followed by *The American Journal of Surgery* (2%, n=5). All other journals were featured 4 or fewer times in our search. Among the 250 studies for which impact factors were available, nearly half (49%, n=122) were published in low-ranked journals as opposed to 32% (n=80) in mid-ranked and 19% (n=48) in top-ranked journals.

The sociodemographic variables across which disparities were studied are displayed in Fig 2. 67% (n=175) of studies covered racial or ethnic disparities. Among these, 56% (n=98) focused exclusively on race/ethnicity, while 15% (n=26) investigated disparities across one additional factor, and 29% (n=51) across two or more additional factors. 10% of studies, including all employment-related studies, described only sex or gender disparities. Studies investigating disparities relating to geography (3%, n=9), rurality (3%, n=9), socioeconomic status (3%, n=8), disability (1.5%, n=4), insurance (1.5%, n=4), and English proficiency (0.8%, n=2) were less frequent. 22 (8%) papers described disparities that fell outside these categories.

**Fig 2.**
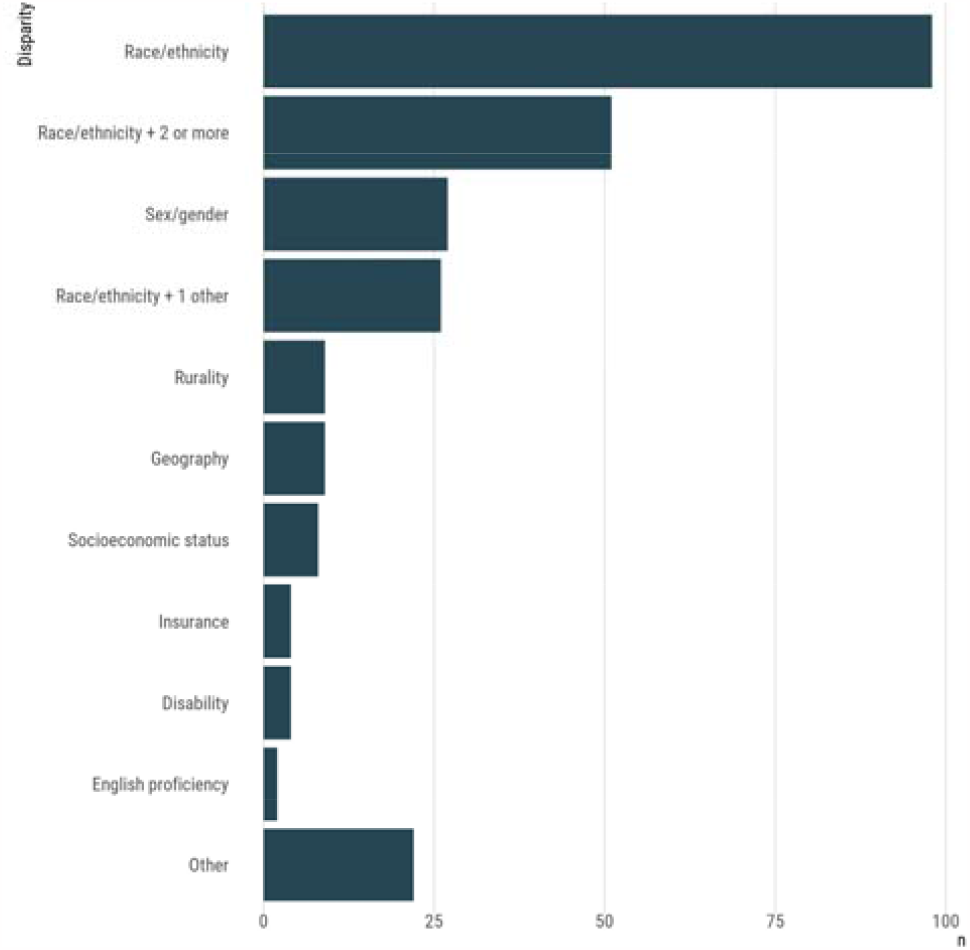
Variables examined in disparity studies

217 of 260 studies provided subjects’ mean or median ages or data from which weighted mean ages could be approximated (Fig 3). Subjects featured in disparity studies tended to be adults, with 71% (n=154) of studies having mean/median ages of participants between 40 and 80. The largest age category was 60 to 70 (29%, n=63), followed by 50 to 60 (17%, n=36) and 40 to 50 (15%, n=32). The mean/median age of participants was 10 or below in 11% (n=23) of studies and between 10 and 20 in 9% (n=19) of studies. Few studies had mean/median ages of subjects between 20 and 30 (3%, n=6), 30 and 40 (5%, n=11), or 80 and 90 (2%, n=4).

**Fig 3.**
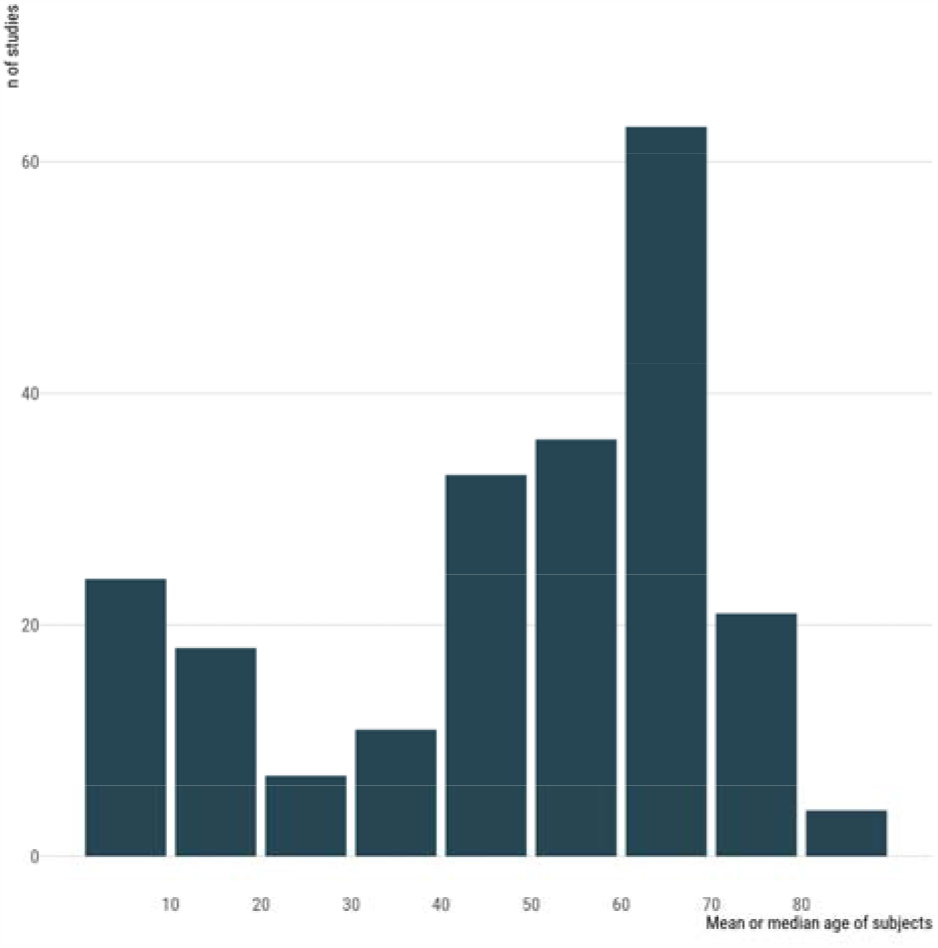
Mean or median ages of subjects in disparity studies

Ages were also grouped into youth (ages 0-17), young adult (ages 18-30), adult (ages 31-64), and older adult (ages 65+) categories. For all except one sociodemographic disparity category (rurality), mean/median ages skewed towards adults and older adults (Fig 4). Young adults were the least frequently featured group in each category. Studies on disparities based on disability, English proficiency, and insurance primarily focused only on adults or older adults. Among studies on racial or ethnic disparities (with or without one or more additional variables), 22% (n=35) focused on youth, 4% (n=6) on young adults, 49% (n=76) on adults, and 25% (n=39) on older adults.

**Fig 4.**
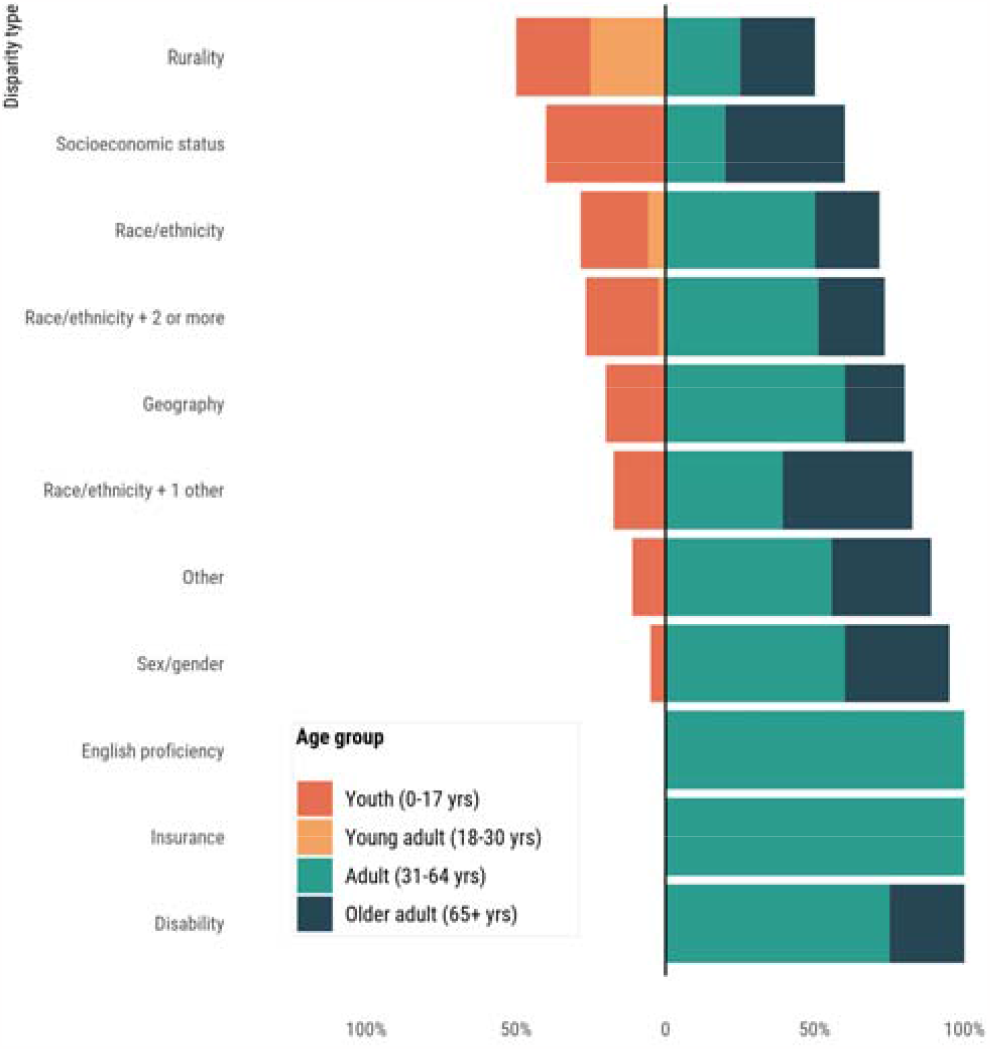
Ages of study subjects by disparity type

Ages of study subjects varied based on the condition under investigation (Fig 5). For most conditions, the vast majority of studies included mainly adult or older adult participants. All studies on cancer, HIV, liver, ophthalmologic, or thyroid/parathyroid-related conditions had mean/median ages of subjects above 30. Young adults were only featured in studies on substance use/psychiatric, obstetrics/gynecologic, or orthopedic conditions. The majority of studies in only three categories—autism/intellectual disability, general primary care/surgical, and pulmonary conditions—focused primarily on pediatric subjects.

**Fig 5.**
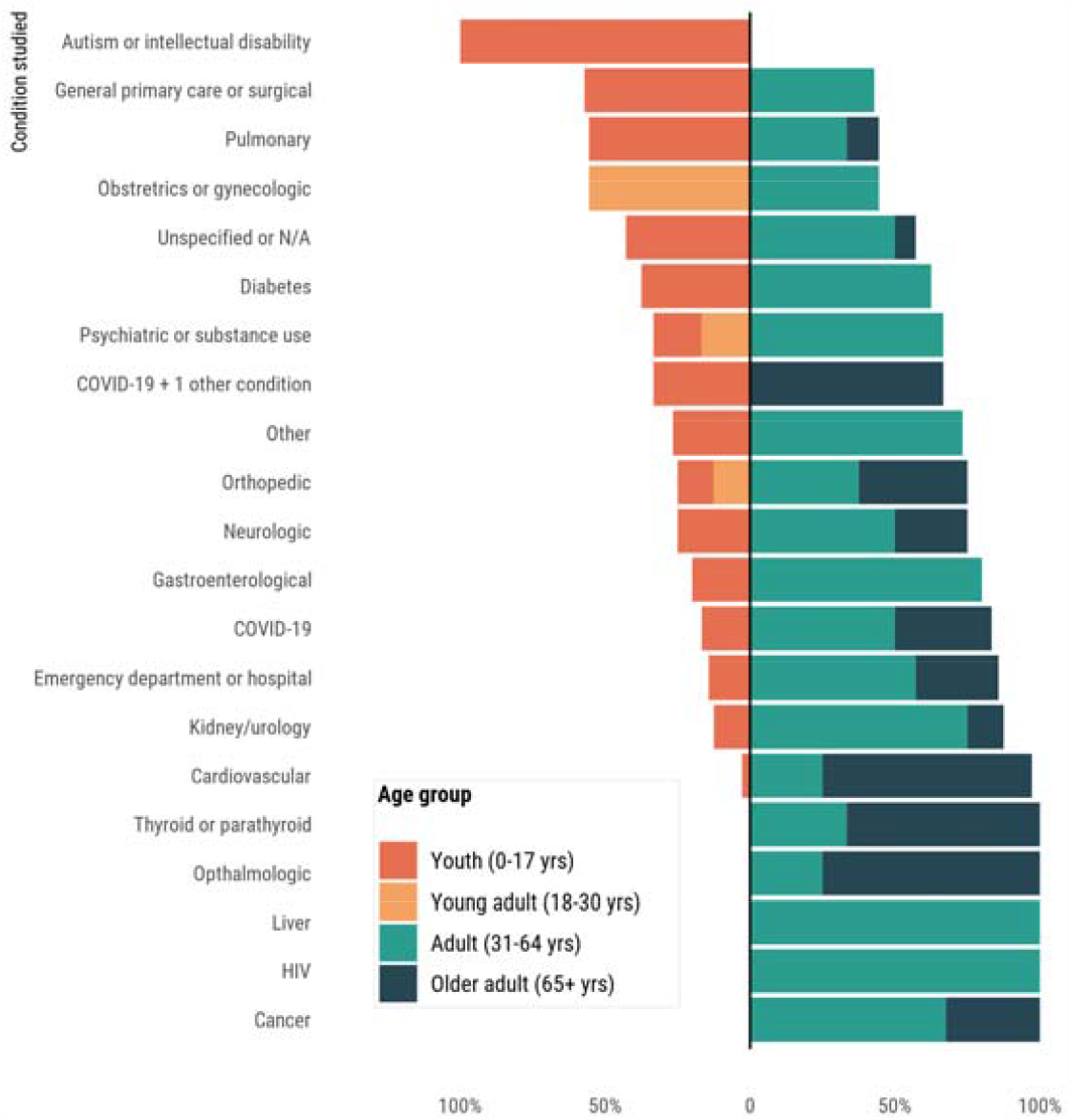
Ages of study subjects by condition studied

## Discussion

Our analysis of 260 research articles on disparities reveals patterns, trends, and gaps in the field. The vast majority of studies focused on health outcomes (95%). When health outcomes were examined, they were disproportionately studied in older ages. The three highest categories of age examined were 60 to 70, 50 to 60, and 40 to 50 years. This suggests an opportunity and need to identify differences in health status at younger ages.

While the vast majority of studies covered health disparities, 9 of 260 studies investigated employment-related disparities among physicians, with 6 focusing on surgical or interventional specialties. These studies characterized disparities in industry payments, award reception, compensation, caregiver responsibilities, autonomy, and mentorship between male and female physicians in various stages of training, establishing that gender disparities are pervasive in medicine. None of the 9 studies explored employment disparities by variables other than gender. It is vital to study career disparities by race, ethnicity, sexual orientation, disability status, and childhood socioeconomic status, as these factors have been well-documented sources of career discrimination in other fields and similar disparities may exist in medicine. Studies on disparities in education were the least common, with just two studies investigating differences in school disciplinary action by race. Such studies are less commonly published in health-related journals and therefore not as likely to appear in a PubMed search.

Cancer was the most frequently studied health condition in our cohort, with 47 studies investigating disparities in cancer incidence, outcomes, treatment, and screening. Among studies focusing on a single cancer type, the mostly frequently studied malignancy was breast cancer, followed by lung and prostate cancers. A 2016 analysis of the 100 most cited papers in health care disparities similarly found cancer to be the top category studied.[6] The popularity of research on cancer and cardiovascular conditions, which together comprised 34% of health-related studies, is expected given that they are among the top causes of disability adjusted life years (DALY) in the United States. Our analysis suggests that drug use disorders and dementia (each <2% of health-related articles) are underrepresented in the field of health disparities despite being leading contributors to DALYs.

The distribution of mean/median ages of subjects included in disparities research reveals a skew towards middle-aged and older adults, with the most commonly studied population aged between 60 and 70 years. Almost all studies on cancer and cardiovascular disease, the most frequently investigated conditions, included primarily adult subjects. Only one-fifth of studies included participants with mean/median ages that were below 20, highlighting the lesser popularity of pediatric disparity research compared to adult disparity research. Yet, many disparities originate in childhood and studies on pediatric disparities can identify groups at higher risk of conditions that present later in life. For instance, Groot et al. studied ethnic disparities in liver fat accumulation in school-age children and identified groups at increased risk of nonalcoholic fatty liver disease.[8] Moreover, ethnic disparities in liver fat appeared to be driven by preventable lifestyle and socioeconomic factors. By targeting at-risk groups before disease develops, interventional approaches may narrow gaps in adult metabolic, cardiovascular, and liver disease incidences between ethnic groups. Another study by Selvaraju et al. compared racial differences in telomere length, a marker of aging, finding that African American children had longer telomeres than European American children.[9] As telomere length is affected by social stress, these findings lend insight into racial differences in susceptibility to cardiovascular disease and can guide early prevention efforts.

Our analysis identified young adults to be the least frequently studied age group in disparities research. Only 3% of studies had mean/median ages of subjects between 20 and 30, and the majority of these investigated disparities in maternal health. Moreover, no studies on disparities in sex/gender, geography, socioeconomic status, disability, insurance, or English proficiency had mean/median ages of subjects in the young adult range. The underrepresentation of young adults in disparities research is multifactorial, likely due to difficulty recruiting research participants, lack of health guidelines for young adults, and the fact that rates of health care utilization are lowest among this population. However, studying disparities in young adults is crucial, as early adulthood represents a unique developmental period that is distinct from childhood, adolescence, and later adulthood.[10] Many conditions like diabetes, coronary artery disease, and autoimmune disease can manifest in early stages in this age range. Young adulthood may thus be a critical time period for implementing interventions to prevent or reverse the development of disease before progression is irreversible. In addition, certain mental health disorders, substance abuse, injuries, and sexually transmitted infections reach their peak in early adulthood.

While documenting disparities is an essential first step, evidence-based interventional approaches are required to bridge differences in outcomes between groups. Only a few studies in our cohort went beyond analyzing data and tested interventions aimed to dismantle health-related disparities. For instance, Cykert et al. designed an intervention for cancer patients that utilized a real-time registry with a navigator and clinical feedback, demonstrating that it narrowed Black-White disparities in cancer treatment completion.[11] Charlot et al. implemented an antiracism intervention using a warning system and race-specific feedback on treatment rates that was found to reduce racial gaps in timely lung cancer surgery.[12] Further exploration of disparity reduction strategies such as policy changes, patient and provider education programs, and community involvement initiatives is needed. A more translational approach to disparities research may guide structural interventions to reduce disparities and yield real-world improvements in health outcomes.

### Strengths and limitations

Our analysis has at least two strengths. By using broad search terms, we were able to identify articles covering disparities in education or employment in addition to articles on health and healthcare. We also categorized studies based on mean or median ages of subjects, manually estimating them when exact values were not provided. This enabled us to identify age groups that are underrepresented in disparities research (for instance, young adults), which may have been missed if studies were more broadly classified as adult or pediatric. There are at least two limitations to our work. A literature review through PubMed likely yields mostly health-related studies and fewer educational and employment-related studies. Using the search term “disparities” rather than “inequalities” may favor studies on health outcomes, patient characteristics, or access to health services over studies on health status.[1] In addition, some studies did not provide any data on ages of participants or provided age data as brackets rather than single mean or median values. In the latter case, we estimated the weighted mean age of subjects and approximated upper and lower bounds if necessary. In some cases, this required us to make assumptions about the lowest or highest ages of study participants as described further in the methods. Some age brackets provided were broad (e.g., 40-60) and estimates of mean may consequently be less accurate.

## Conclusion

Our systematic analysis of research articles on disparities found that the majority of studies investigated health or healthcare-related disparities, of which more than a third covered cancer and cardiovascular disease. The majority of papers (67%) covered racial or ethnic disparities. Studies assessing physician career outcomes were three times more common than studies assessing educational outcomes in children. Physician career outcomes were only examined for evidence of gender inequity. Studies on disparities focused primarily on adult populations, with 71% of studies having mean or median ages of subjects between the ages of 40 and 80, compared to only 19% below the age of 20. 20 to 30-year-olds were the least frequently studied age group (3%). Our examination reveals persistent gaps in the disparities literature. A greater focus on disparities in childhood and early adulthood may lead to early intervention strategies to improve long-term health and reduce disparities manifesting later in life.

## Data Availability

All data produced in the present study are available upon reasonable request to the authors

## Declarations

### Funding

There were no funds, grants, or other support provided for this study.

### Conflicts of Interest/Competing interests

Vinay Prasad’s disclosures: (Research funding) Arnold Ventures (Royalties) Johns Hopkins Press, Medscape, and MedPage (Honoraria) Grand Rounds/lectures from universities, medical centers, non-profits, and professional societies. (Consulting) UnitedHealthcare and OptumRX. (Other) Plenary Session podcast has Patreon backers, YouTube, and Substack. All other authors have no financial nor non-financial conflicts of interest to report.

### Availability of data and material

The data that support the findings of this study are openly available in PubMed.

### Code availability

Not applicable

### Author contributions

All authors contributed to study conception and design. Material preparation, data collection and analysis were performed by Anushka Walia, Alyson Haslam, and Milos Milijković. Data presentation was performed by Jordan Tuia. The first draft of the manuscript was written by Anushka Walia and all authors commented on previous versions of the manuscript. All authors read and approved the manuscript.

### Ethics approval

Institutional review board approval was not required because data were publicly available and study does not use patient-specific data.

### Consent to participate

Not applicable since this research did not use individual participants.

### Consent for publication

Not applicable since this research did not use individual participants.

## Notes

### Competing Interest Statement

Vinay Prasad Disclosures: (Research funding) Arnold Ventures (Royalties) Johns Hopkins Press, Medscape, and MedPage (Honoraria) Grand Rounds/lectures from universities, medical centers, non-profits, and professional societies. (Consulting) UnitedHealthcare and OptumRX. (Other) Plenary Session podcast has Patreon backers, YouTube, and Substack. All other authors have no financial nor non-financial conflicts of interest to report.

### Funding Statement

This study did not receive any funding

### Summary of Updates

Fixed images and changed abstract so it was not formatted.

